# Reassessing Fragility: A Comparative Analysis of the Fragility Index With the Relative Risk Index

**DOI:** 10.1101/2023.10.04.23296567

**Authors:** Thomas F. Heston

## Abstract

**Background:** In biostatistics, assessing the fragility of research findings is crucial for understanding their clinical significance. This study focuses on the fragility index, unit fragility index, and relative risk index as measures to evaluate statistical fragility. The relative risk index quantifies the deviation of observed findings from therapeutic equivalence. In contrast, the fragility indices assess the susceptibility of p-values to change significance with minor alterations in outcomes within a 2×2 contingency table. While the fragility indices have intuitive appeal and have been widely applied, their behavior across a wide range of contingency tables has not been rigorously evaluated.

**Methods:** Using a Python software program, a simulation approach was employed to generate random 2×2 contingency tables. All tables under consideration exhibited p-values < 0.05 according to Fisher’s exact test. Subsequently, the fragility indices and the relative risk index were calculated. To account for sample size variations, fragility, and risk quotients were also calculated. A correlation matrix assessed the collinearity between each metric and the p-value.

**Results:** The analysis included 2,000 contingency tables with cell counts ranging from 20 to 480. Notably, the formulas for calculating the fragility indices encountered limitations when cell counts approached zero or duplicate cell counts hindered standardized application. The correlation coefficients with p-values were as follows: unit fragility index (-0.806), fragility index (-0.802), fragility quotient (-0.715), unit fragility quotient (-0.695), relative risk index (-0.403), and relative risk quotient (-0.261).

**Conclusion:** Compared with the relative risk index and quotient, in the context of p-values < 0.05, the fragility indices and their quotients exhibited stronger correlations. This implies that the fragility indices offer limited additional information beyond the p-value alone. In contrast, the relative risk index displays relative independence, suggesting that it provides meaningful insights into statistical fragility by assessing how far observed findings deviate from therapeutic equivalence, regardless of the p-value.

## Introduction

Statistical fragility refers to the susceptibility of research findings to change from statistically significant to nonsignificant with only minor alterations in the data (1). It is a critical issue in biomedical research because findings that hinge on small numbers of events may not be reproducible. Spurious correlations from small studies are well documented (2), and approximately half of clinical trials are not replicable (3). This undermines the reliability of published findings and wastes resources when initial results fail to translate into patient benefit.

One proposed solution is the unit fragility index (UFI) (1). This looks at the effect of a single unit alteration in each cell of a 2 x 2 contingency table such that the marginal totals remained fixed. This was expanded upon by Walsh et al. (4) to define the fragility index (FI) as the minimal alteration of just two cells of a 2×2 contingency table that would change it from significant to nonsignificant (4). The FI was specifically designed to evaluate the fragility of statistically significant 2×2 contingency tables where the p-value was < 0.05.

The FI thus provides an intuitive stress test for assessing result reproducibility. For instance, an FI of three suggests only three miscategorized events could flip the p-value of a 2×2 contingency table from significant to insignificant. Also, comparing the FI to patient dropout rates adds meaningful information, indicating that the results are highly fragile if the FI is lower than the number of participants lost to follow-up. Reviews across research disciplines consistently have found low fragility indices, suggesting that statistical fragility is a major cause of the reproducibility crisis (5).

However, the FI has limitations. It strongly depends upon the sample size; as the sample size grows, FI increases, incorrectly implying greater robustness (6). Although the fragility quotient (FQ) attempts to address this issue by dividing the FI by sample size (7), it hasn’t been rigorously tested. There also are problems with calculating the FI at low sample sizes, leading to the need for alternative methods of calculating the FI, such as quantifying the minimum change across the entire contingency table (8). There are also concerns that it overemphasizes p-values and dichotomizes results.

However, the p-value approach to analyzing the meaning of a 2×2 contingency table isn’t necessarily the best method for clinically applying the results. In clinical practice, often there is incomplete information regarding which treatment or which test is optimal. In these cases the relative risk can help guide clinical management. The p-value evaluates how far the contingency table varies from random chance, and the FI helps determine how confident we can be in that finding. The relative risk, on the other hand, doesn’t evaluate deviation from random chance. It quantifies the risk of one treatment over another. When the relative risk is high in the sample studied, one treatment is much better or worse than the other. The two treatments are equivalent and therapeutically neutral when the relative risk equals one. The relative risk index is a novel metric that quantifies how far a contingency table is from therapeutic neutrality. It the average of the absolute value of the observed minus expected value for each cell in the table. In a 2×2 contingency table it equals |(ad-bc)|/N where a, b, c, and d represent the observed outcomes in each of the four cells and N equals the sample size.

In this study, the behavior of the fragility indices and their quotients are evaluated using simulated randomly generated 2 x 2 contingency tables associated with a statistically significant p-value (<0.05). These measures of fragility are then compared with the average of the contingency table’s residuals, referred to here as the relative risk index (RRI). The RRI was also divided by the sample size to give a risk quotient (RQ). Correlations with the p-value were then performed to evaluate how much additional information each metric provided beyond the p-value. High correlation coefficients suggest that the metric provides little information beyond the p-value, whereas a low correlation coefficient would suggest the opposite.

A preliminary draft of this manuscript was published on the Authorea preprint server on August 25, 2023 (9).

## Methods

### Random table generation

Python was used to generate random 2×2 contingency tables. The cells were labeled in standard fashion (Table 1) throughout the Python code. Additionally, throughout this manuscript, cells are referred to in this standardized manner. Specifically, Table 1 is referred to as (a, b, c, d). Individual cell values ranged from 20 to 480. Because the FI was specifically designed to provide further insight into the meaning of *statistically significant* 2×2 contingency tables, only tables associated with Fisher’s exact p-value below 0.05 were included in the study sample. The data and code are publicly available in the Zenodo repository (10).

**Table 1.** Standard nomenclature for a 2×2 contingency table.

| **Treatment Groups** | **Outcome A** | **Outcome B** | **Marginal Totals** |
| --- | --- | --- | --- |
| **Group A** | a | b | a + b |
| **Group B** | c | d | c + d |
| **Marginal Totals** | a + c | b + d | N |

**Legend:** a, b, c, and d represent the number of subjects in each treatment group experiencing either Outcome A or Outcome B. N is the total sample size.

### Calculation of the fragility index and quotient

The FI was calculated based on the method described by Walsh et al. (4). The cell with the lowest number of outcomes was incremented by one until the statistical significance of the table, as calculated by Fisher’s exact test, flipped from less than 0.05 to greater than 0.05. Marginal totals for the rows, but not the columns, are kept fixed (Table 2). The FQ was calculated as the FI divided by the sample size (N).

**Table 2.** Calculation of the fragility index.

| **Treatment Groups** | **Outcome A** | **Outcome B** | **Marginal Totals** |
| --- | --- | --- | --- |
| **Group A** | $a + f$ | $b - f$ | $a + b$ |
| **Group B** | $c$ | $d$ | $c + d$ |
| **Marginal Totals** | $a + c + f$ | $b + d - f$ | $N$ |

**Legend:** in the scenario where cell "a" has the fewest events, the fragility index was calculated as the smallest integer value for "f" causing Fisher's exact test to flip from significant to insignificant.

### Calculation of the unit fragility index and quotient

The UFI was calculated based on the method described by Feinstein (1). The cell with the lowest number of outcomes was incremented by one until the statistical significance of the table, as calculated by Fisher’s exact test, flipped from less than 0.05 to greater than 0.05. Marginal totals are kept fixed (Table 3). The unit fragility quotient (UFQ) was calculated as the UFI / N.

**Table 3.** Calculation of the unit fragility index.

| **Treatment Groups** | **Outcome A** | **Outcome B** | **Marginal Totals** |
| --- | --- | --- | --- |
| **Group A** | a + f | b - f | a + b |
| **Group B** | c - f | d + f | c + d |
| **Marginal Totals** | a + c | b + d | N |

**Legend:** in the scenario where Outcome A has the fewest events, the fragility index was the smallest integer (f) causing Fisher's exact test to flip from significant to insignificant.

### Calculation of the relative risk index and quotient

As previously stated, the RRI is the average residual of the table. The residual is the difference between the observed count in a cell and the expected count for that cell. In the case of 2×2 contingency tables, the RRI equals the |(ad-bc)|/N. After the RRI modifies each cell, the positive predictive values for each treatment group become equal, and the relative risk equals one. This is the point of therapeutic neutrality for each of the two treatments. The RQ is the RRI / N. The RRI and the RQ are derived from observed values, do not quantify the impact of case misclassifications, and do not rely on a dichotomous change in statistical significance. The marginal totals are kept equal (Table 4).

**Table 4.** Calculation of the relative risk index for 2×2 contingency tables.

| Treatment Groups | Outcome A | Outcome B | Marginal Totals |
| --- | --- | --- | --- |
| Group A | $a + r$ | $b - r$ | $a + b$ |
| Group B | $c - r$ | $d + r$ | $c + d$ |
| Marginal Totals | $a + c$ | $b + d$ | N |

**Legend:** in the scenario where the positive predictive value of Group A is less than that for Group B, the RRI (“r”) is added to cell “a” and the remaining three cells are modified such that the marginal totals are kept equal. Note that the RRI is a real number and not limited to integer values. In 2x2 contingency tables, r equals $|ad-bc|/N$ .

### Case examples

In addition to simulated data, two case examples are provided to demonstrate the application of the FI, FQ, UFI, FQ, RRI, and RQ metrics to real research data.

### Statistical analysis

Statistical analyses were performed by IBM SPSS (version 29). The Pearson correlation coefficient was determined to evaluate the relationships of measures of fragility with the observed p-value of the contingency table. Collinearity was considered significant for variance inflation factor (VIF) values over 10. Scatterplots were generated along with the corresponding regression line. The Fisher r to z transformation was used to compare correlation coefficients (11). Because of the large sample size, the robustness index was applied to assess the resultant p-value’s fragility (12).

## Results

There were 2000 contingency tables analyzed for a p-value of 0.001 to 0.05. There were multiple instances where the formulas for the FI and UFI would not converge to a change in the p-value to > 0.05. This necessitated modifying the Python code to discard tables that did not converge. For example, the p-value for the contingency table of (89, 47, 43, 41) will not flip to insignificant using the formula for the FI or the UFI. In this case, the FI and UFI would need to be negative to change the significance of the table, or alternatively, the formula for the FI and UFI would need to be changed. Instead of just *adding* an event to the smallest outcome, the definition would need to be changed to *adding or subtracting* from the smallest outcome (Table 5).

**Table 5.**
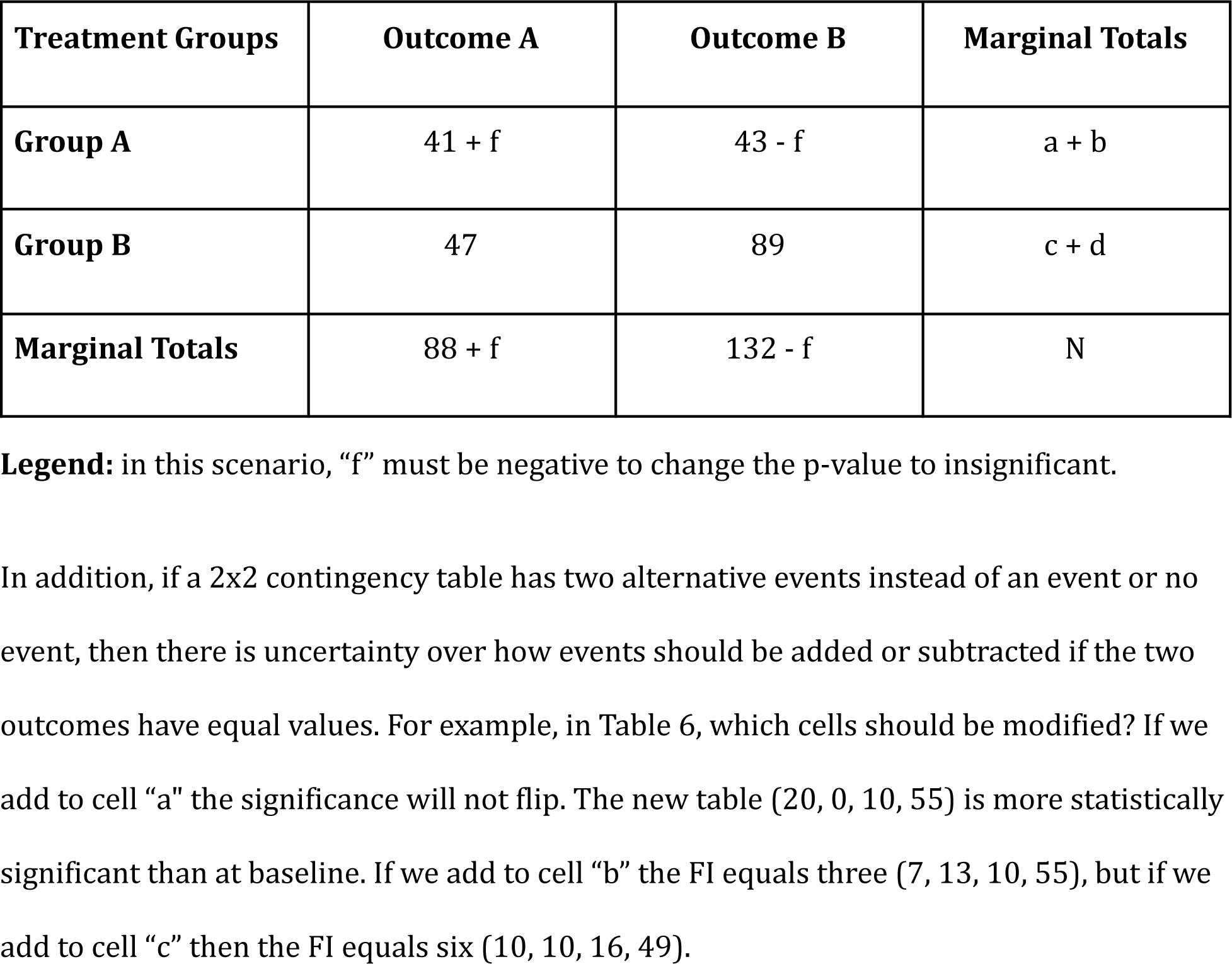
A negative fragility index.

| Treatment Groups | Outcome A | Outcome B | Marginal Totals |
| --- | --- | --- | --- |
| Group A | $41 + f$ | $43 - f$ | $a + b$ |
| Group B | 47 | 89 | $c + d$ |
| Marginal Totals | $88 + f$ | $132 - f$ | N |

**Legend:** in this scenario, “f” must be negative to change the p-value to insignificant.

In addition, if a 2×2 contingency table has two alternative events instead of an event or no event, then there is uncertainty over how events should be added or subtracted if the two outcomes have equal values. For example, in Table 6, which cells should be modified? If we add to cell “a” the significance will not flip. The new table (20, 0, 10, 55) is more statistically significant than at baseline. If we add to cell “b” the FI equals three (7, 13, 10, 55), but if we add to cell “c” then the FI equals six (10, 10, 16, 49).

**Table 6.** Uncertainty of the fragility index.

| **Treatment Groups** | **Outcome A** | **Outcome B** | **Marginal Totals** |
| --- | --- | --- | --- |
| **Group A** | 10 | 10 | 20 |
| **Group B** | 10 | 55 | 65 |
| **Marginal Totals** | 20 | 65 | 85 |

**Legend:** in this scenario, it is unclear which cells should be modified to calculate the fragility index. The fragility index is not constant in this case and will differ depending on the choice of which cells to modify.

Because of these uncertainties of the FI and UFI, all tables were discarded from analysis when a cell incremented to zero or where identical cell counts made it unclear which cell to modify. The Python code was then modified to discard such tables automatically, such that the total number of tables analyzed remained at 2000.

Bivariate correlations were then examined between the p-value and the other variables (FI, FQ, UFI, UFQ, RRI, and RQ). This indicated that FI, FQ, UFI, and UFQ all demonstrated high collinearity with p-value, evidenced by correlation coefficients from -0.695 to -0.806. However, RRI (r = -0.403) and RQ (r = -0.261) had substantially weaker correlations with the p-value (Table 7). After controlling for the other variables, partial correlations between RRI, RQ, and the p-value were also small, at 0.071 and 0.13, respectively.

**Table 7.** Bivariate correlation coefficients.

| Variable | P-Value | FI | FQ | UFI | UFQ | RRI | RQ |
| --- | --- | --- | --- | --- | --- | --- | --- |
| P-Value | 1 | -.802 | -.715 | -.806 | -.695 | -.403 | -.261 |
| FI | -.802 | 1 | .566 | .962 | .494 | .704 | .011 (ns) |
| FQ | -.715 | .566 | 1 | .503* | .945 | -.029 | .718 |
| UFI | -.806 | .962 | .503 | 1 | .503 | .704 | -.021 (ns) |
| UFQ | -.695 | .494 | .945 | .503 | 1 | -.106 | .768 |
| RRI | -.403 | .704 | -.029 (ns) | .704 | -.106 | 1 | -.571 |
| RQ | -.261 | .011(ns) | .718 | -.021 (ns) | .768 | -.571 | 1 |

**Legend:** all correlations are significant at the 0.01 level (2-tailed) except where noted by “ns” (nonsignificant). FI = fragility index, FQ = fragility quotient, UFI = unit fragility index, UFQ = unity fragility quotient, RRI = relative risk index, RQ = risk quotient.

A regression of the scatterplot of the FQ with the p-value showed an R-square value of 0.51, indicating that 51% of the variation in the FQ could be explained by the p-value (Figure 1). The regression of the scatterplot of the RQ with the p-value showed a significantly lower R-square value of 0.068, indicating relative independence of the RQ from the p-value, with only 6.8% of the variation in the RQ explainable by the p-value (Figure 2). The FQ was highly correlated with the UFQ, showing an R-square value of 0.893, indicating that 89% of the variation in the FQ could be explained by the UFQ (Figure 3).

**Figure 1.**
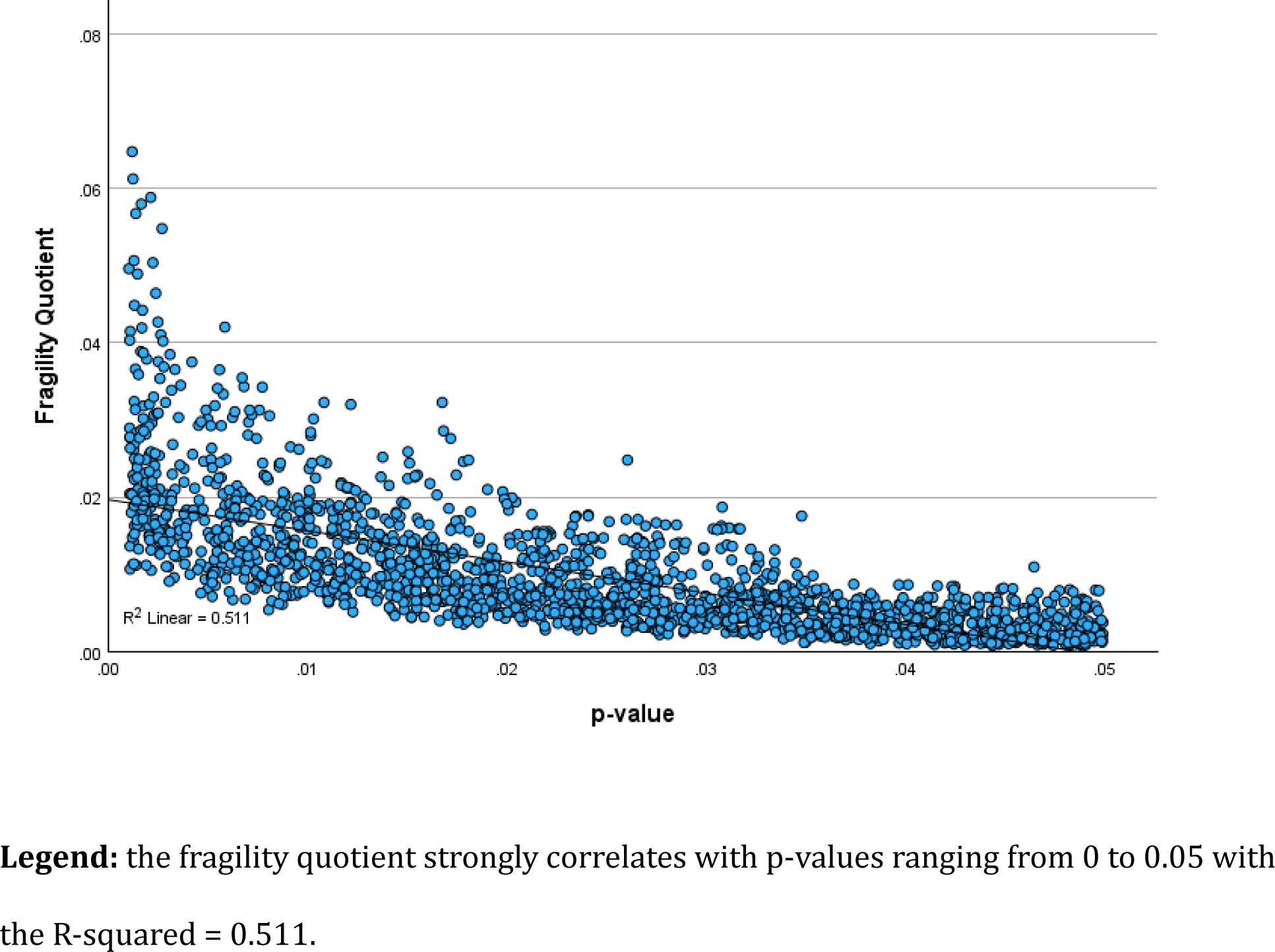
Scatterplot of the FQ with the p-value.

**Figure 2.**
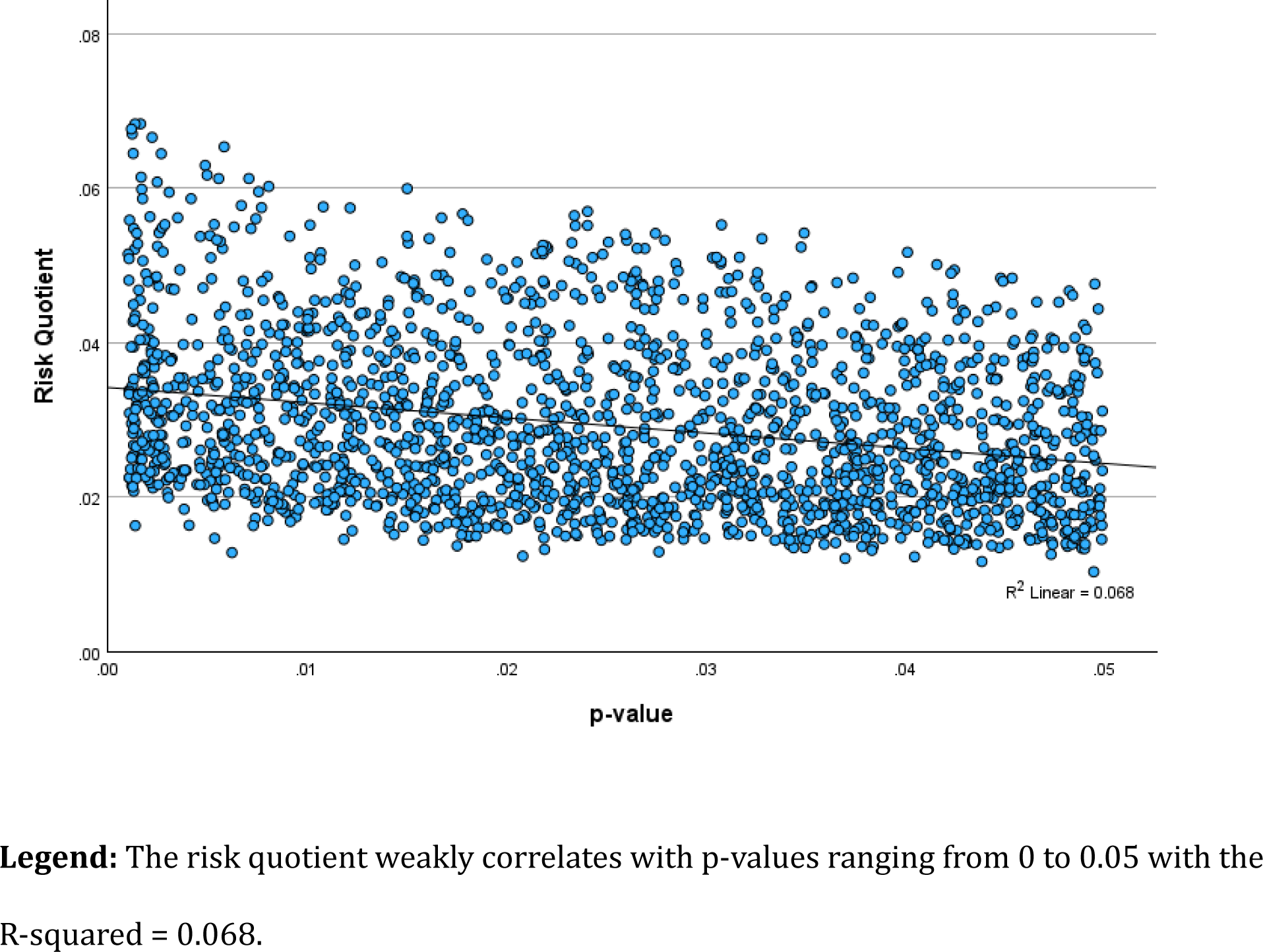
Scatterplot of the RQ with the p-value.

**Figure 3.**
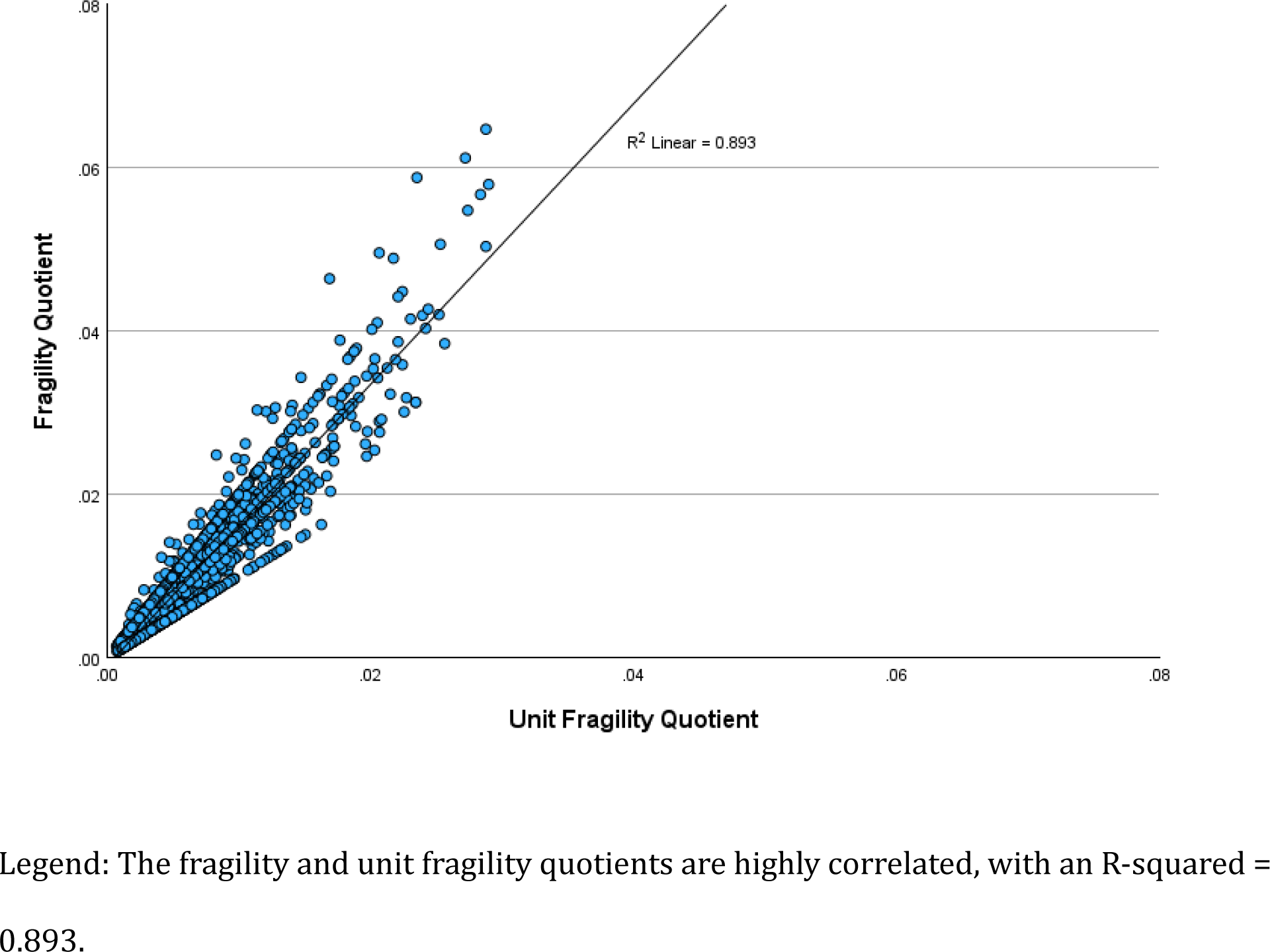
Scatterplot of the FQ with the UFQ.

The difference when comparing the correlation coefficient of the FQ with the p-value (-.715) compared to the RQ with the p-value (-.261) was statistically significant (two-tailed p < 0.001) for the sample size of 2000. This comparison would remain significant even if the sample size were only 23, giving a robustness index of 87 (2000/23), indicating a highly robust difference in these correlation coefficients.

Multiple linear regression was conducted using the enter method to predict the p-value. The predictor variables entered into the model were FI, FQ, UFI, UFQ, RRI, and RQ.

Examining collinearity statistics revealed high multicollinearity between four predictors, as indicated by variance inflation factors (VIFs) exceeding the commonly used cutoff of 10. Specifically, FI had a VIF of 46, FQ had a VIF of 31, UFI had a VIF of 47, and UFQ had a VIF of 38, indicating significant collinearity between these four variables. In contrast, RRI and RQ demonstrated acceptable VIFs of 6 and 7, respectively, indicating these variables were not collinear with the other predictors in the model.

### Case studies

A study of minimally invasive surfactant therapy (MIST) in preterm infants found that hospitalization in the first two years was 25.1% in the MIST group and 38.2% in the control group (13). The 2×2 contingency table was (49, 146, 78, 126), and by Fisher’s exact the p-value is 0.0053. The FI is seven (56, 139, 78, 126) with the FQ = 0.018. The UFI is four (53, 142, 74, 130) and UFQ = 0.010. The RRI = |(49 * 126 - 146 * 78)/399| = 13.07 and the RQ = 0.0328. Although the p-value is highly significant, the RQ is in the moderate category of 0.02 to 0.04. The p-value, FI, FQ, UFI, and UFQ all suggest highly robust findings. Looking at Figure 2, for a p-value of approximately 0.005, an RQ of 0.0328 indicates only mild to moderate robustness, suggesting that some skepticism should be taken into consideration when making clinical decisions on this topic. These findings suggest that the RQ appears to provide information independent of the p-value.

A study of video intervention and documentation of goals of care found that documentation was greater in the intervention group than in the control group (14). The 2×2 contingency table was (3744, 2279, 2396, 2383), with a p-value of < 0.001 by Fisher’s exact. The FI is 610, the FQ is 0.056, the UFI is 271, and the UFQ is 0.025. The RRI = 320.4 and the RQ = 0.028. In this case, the p-value, FI, FQ, UFI, and UFQ are all consistent with highly robust findings. Looking at Figure 2, an RQ of 0.028 is in the lower range for a p-value of < 0.001, consistent with at least moderately decreased robustness.

## Discussion

The FI and UFI demonstrated several problems, making it impossible to apply their formulas uniformly. They cannot change the significance of some tables unless they take on negative values. If some cell counts are equal, the FI and UFI will differ depending upon an arbitrary choice of which cells to modify. The high collinearity among the p-value and FI, FQ, UFI, and UFQ signifies these four variables are redundant with the p-value and add little additional useful information. On the other hand, the lack of collinearity between RRI, RQ, and the p-value suggests that RRI and RQ contain meaningful information beyond what the p-value provides; their minimal correlations imply they measure something substantially different. Given their non-redundancy with the p-value or the other study variables, RRI and RQ likely contribute unique and useful information to complement the p-value when interpreting study findings. The RRI and RQ provided non-redundant information beyond what is contained in the p-value. The weak bivariate and partial correlations between RRI, RQ, and the p-value indicate that RRI and RQ capture information distinct from what is contained within the p-value. Although only two case studies from the literature were analyzed, they did show relative independence of the RQ from the p-value and the other metrics of fragility.

The problems with calculating the FI have been recognized previously when attempting to integrate this metric into the R statistical package. One solution to address the issue of a cell value going to zero, making the application of Fisher’s exact test impossible, has been to substitute the chi-square test in these instances (15). Another proposed solution is to adjust the formula for FI, use absolute values, and when the FI cannot be calculated, simply set the FI to equal “not available” (8). While these solutions partially address calculation problems, they do not address the underlying issue of the FI and UFI being dependent upon an arbitrary p-value-based statistical significance threshold (16).

The strong correlations between the FI, UFI, FQ, and UFQ with the p-value indicate that they provide redundant information. Correlation coefficients greater than |0.7|, as seen in our analysis, are problematic and can distort several statistical data analyses (17). Our findings confirm previous simulations showing a strong correlation between the FI and p-value (6). However, our analysis extends this finding to the FQ, UFI, and UFQ. Since these metrics of fragility all hinge upon flipping the p-value from significant to insignificant, it is not surprising that they all strongly correlate with the p-value. Nevertheless, our data suggests that if any of these metrics are used in addition to the p-value, the most informative ones correct for sample size, specifically the FQ or the UFQ.

An important finding of our simulation study is that the RRI and the RQ are only weakly correlated with p-values ranging from 0 to 0.05. In addition to the weak correlation coefficients, VIF analysis showed low collinearity, and partial correlation after controlling for the other variables remained weak. These findings confirm that looking at relative risks rather than a flipping of p-value significance may be a better way to evaluate a study’s fragility. The RRI and RQ both appear to provide meaningful information that goes beyond what is contained in the p-value. Perhaps the best demonstration of this is the scatterplot of the RQ versus the p-value (Figure 2). This suggests that for any fixed p-value < 0.05, an RQ of 0.02 or less indicates relatively fragile findings, an RQ of 0.02 to 0.04 indicates moderate robustness and an RQ of over 0.04 suggests robust findings.

The clinical implications of these findings are meaningful. Rather than research findings focusing on an arbitrary p-value and the fragility of that arbitrary value, the RRI focuses on relative risks, the most important factor in clinical decision-making. Ultimately, clinicians want to know the clinical impact of a research study. This frequently boils down to one simple question: is the new treatment more likely than not to be of net benefit? This “more likely than not” is relative risk.

A high RRI, well above one, indicates that the treatment in question is much more likely than not to be of benefit. Even a low RRI will indicate which treatment is more likely than not to be of benefit. Alternatively, an RRI of one indicates treatment neutrality, at which neither treatment is more likely than the other to benefit the patient. If a new treatment has a low RRI over conventional treatment, then clinicians would be wise to be skeptical of whether or not the new treatment is beneficial, even if the p-value of the study is statistically significant.

The proposal of confidence intervals replacing p-values is also problematic because they are based on populations, not individuals. While the standard deviation is useful when determining what is best for an individual patient, confidence intervals only address large population differences evaluated in a research study. While useful in this setting, they have less value in clinical practice. For example, the formula for calculating a 95% confidence interval of a sample mean equals:

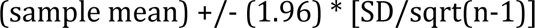

where SD equals the standard deviation of the sample, and n is the sample size. Thus, for large studies, the divisor, the square root of n-1, will dramatically decrease the size of the standard error. The result is that studies with large sample sizes are more likely than studies with small sample sizes to be statistically significant. But when it comes to individual patient management, standard deviations, not standard errors, are most relevant. The net effect is that p-values and the closely associated fragility indices FI, FQ, UFI, and UFQ have benefits when analyzing populations but not so much individual patients.

The RRI and RQ, on the other hand, are particularly useful to clinicians responsible for the care of individual patients. A good, skeptical clinician will always question the fragility of a study. With the RRI and RQ, clinicians can quantify just how large or small a change in outcomes is required to make the new treatment or test of zero benefit. The endpoint isn’t a flip in an arbitrary p-value cutoff but treatment neutrality. For example, if the RQ is very low, clinicians may want to stay with an older, more proven therapy even if the p-value of the study is statistically significant. If the RQ is high, clinicians and their patients can have greater confidence that a newer, more expensive treatment or test may be worth it.

This study has several important limitations. First, the data consisted of randomly generated 2×2 contingency tables rather than real-world data from clinical trials or observational studies. While simulated data is useful for initial testing, real-world data is needed to determine if these findings translate to actual research scenarios.

Second, the analysis was restricted to tables with p-values ranging from 0 to 0.05. This narrow range was chosen because the fragility index as originally proposed was designed for significant p-values only, but evaluating a wider range of p-values could provide further insights into the behavior of the RRI and RQ. As the p-value increases, the RRI is anticipated to steadily taper towards a value of one, but the FI is anticipated to follow a curve with high values at each extreme with the lowest FI values around a p-value of 0.05. Nevertheless, their performance with non-significant p-values remains not fully characterized.

Third, the RRI and RQ have not yet been sufficiently tested with real data across diverse research disciplines. Demonstrating their practical utility will require rigorous testing with published findings. This is particularly important for validating the proposed categories of robustness based on RQ values.

Fourth, it is unknown how the RRI and RQ will perform with more complex study designs beyond simple 2×2 contingency tables. Testing with data from studies with multiple arms, covariates, and more advanced statistical analyses is needed. However, since the RRI is simply the average residual, i.e. the average of the expected minus observed value for each cell, it likely will be easily extended to larger contingency tables. However, it is not clear how the RQ score will behave, and if the proposed categories of robustness will hold.

Finally, while this initial investigation focused on statistical fragility, the RRI and RQ may have applications beyond this context that require further exploration. Determining their value for assessing clinical relevance or predictive modeling warrants future research.

In summary, this study provides early evidence that the RRI and RQ may provide useful complements to p-values and fragility indices, but considerable additional research with real-world data is needed to substantiate their utility across different statistical scenarios and research disciplines. Testing their application for supporting clinical decision-making is particularly important.

## Conclusion

This study demonstrates that the RRI and RQ provide measures of statistical fragility that are less dependent on p-values than the FI, FQ, UFI, and UFQ. The RRI and RQ also do not run into the calculation problems of the fragility indices. The weak correlation of RRI and RQ with p-values indicates that they provide different information beyond what is captured by p-values alone. The RRI and RQ add nuance beyond significance testing for clinicians evaluating treatments and diagnostic tests. Overall, this initial investigation of the RRI and RQ as alternatives to the FI, FQ, UFI, and UFQ shows promise for providing a metric of fragility that is less reliant on arbitrary p-value cutoffs.

## Data Availability

All data produced are available online at the Zenodo repository.

https://www.zenodo.org/record/8408743

## FUNDING INFORMATION

Self-funded, no external funding.

## CONFLICT OF INTERESTS

No competing interests.

## ETHICAL APPROVAL

This study did not involve human or animal research.

## LICENSE

CC BY 4.0

## Notes

### Competing Interest Statement

The authors have declared no competing interest.

### Funding Statement

This study did not receive any funding

## Bibliography

1. Feinstein AR. The unit fragility index: an additional appraisal of “statistical significance” for a contrast of two proportions. J Clin Epidemiol. 1990;43(2):201–9.

2. Ioannidis JPA. Why most published research findings are false. PLoS Med. 2005 Aug 30;2(8):e124.

3. Niven DJ, McCormick TJ, Straus SE, Hemmelgarn BR, Jeffs L, Barnes TRM, et al. Reproducibility of clinical research in critical care: a scoping review. BMC Med. 2018 Feb 21;16(1):26.

4. Walsh M, Srinathan SK, McAuley DF, Mrkobrada M, Levine O, Ribic C, et al. The statistical significance of randomized controlled trial results is frequently fragile: a case for a Fragility Index. J Clin Epidemiol. 2014 Jun;67(6):622–8.

5. Bertaggia L, Baiardo Redaelli M, Lembo R, Sartini C, Cuffaro R, Corrao F, et al. The Fragility Index in peri-operative randomised trials that reported significant mortality effects in adults. Anaesthesia. 2019 Aug;74(8):1057–60.

6. Carter RE, McKie PM, Storlie CB. The Fragility Index: a P-value in sheep’s clothing? Eur Heart J. 2017 Feb 1;38(5):346–8.

7. Ahmed W, Fowler RA, McCredie VA. Does sample size matter when interpreting the fragility index? Crit Care Med. 2016 Nov;44(11):e1142–3.

8. Lin L, Chu H. Assessing and visualizing fragility of clinical results with binary outcomes in R using the fragility package. PLoS ONE. 2022 Jun 1;17(6):e0268754.

9. Heston TF. A Comparative Analysis of Unit Fragility and the Relative Risk Index. Authorea. 2023 Aug 25;

10. Heston TF. Dataset for comparison of the Fragility Index with the Relative Risk Index. 2023 Oct 4;

11. Lowry R. Significance of the Difference Between Two Correlation Coefficients [Internet]. VassarStats. [cited 2023 Sep 30]. Available from: http://vassarstats.net/rdiff.html

12. Heston TF. The robustness index: going beyond statistical significance by quantifying fragility. Cureus. 2023 Aug 30;

13. Dargaville PA, Kamlin COF, Orsini F, Wang X, De Paoli AG, Kanmaz Kutman HG, et al. Two-Year Outcomes After Minimally Invasive Surfactant Therapy in Preterm Infants: Follow-Up of the OPTIMIST-A Randomized Clinical Trial. JAMA. 2023 Sep 11;

14. Volandes AE, Zupanc SN, Lakin JR, Cabral HJ, Burns EA, Carney MT, et al. Video Intervention and Goals-of-Care Documentation in Hospitalized Older Adults: The VIDEO-PCE Randomized Clinical Trial. JAMA Netw Open. 2023 Sep 5;6(9):e2332556.

15. Johnson K. fragilityindex: Repository for the R Package Fragility Index [Internet]. Github. 2017 [cited 2023 Oct 4]. Available from: https://github.com/kippjohnson/fragilityindex

16. Potter GE. Dismantling the Fragility Index: A demonstration of statistical reasoning. Stat Med. 2020 Nov 20;39(26):3720–31.

17. Dormann CF, Elith J, Bacher S, Buchmann C, Carl G, Carré G, et al. Collinearity: a review of methods to deal with it and a simulation study evaluating their performance. Ecography. 2013 Jan;36(1):27–46.

